# Detection of Measles in Texas Wastewater

**DOI:** 10.1101/2025.04.08.25325475

**Authors:** Katherine M. Joseph, Xingwen Chen, Dhvani Parikh, Janelle Rios, Catherine L. Troisi, Michael J. Tisza, Anthony W. Maresso, Blake M. Hanson, Anna Gitter, Jennifer Deegan, Cici X. Bauer, John Balliew, Kristina D. Mena, Eric Boerwinkle, Fuqing Wu

## Abstract

Measles outbreaks continue to pose significant public health challenges globally despite the availability of effective vaccines. In this study, we evaluated wastewater-based surveillance for detection of measles virus during an ongoing outbreak in Texas. Weekly wastewater samples collected from two Texas cities between January 2 and March 17, 2025 were analyzed using multiple RT-PCR assays targeting the nucleoprotein and matrix genes of the measles virus. Viral RNA was detected in multiple days from both cities, with City A showing positives from January 13 and City B from January 6, both predating the first confirmed case in the state on January 23. Sequencing of PCR amplicons confirmed the specificity of detection and phylogenetic analysis using global and U.S. measles genome databases further validated that the viral RNA belonged to the currently circulating genotype D8. Our findings demonstrate that wastewater surveillance can provide early evidence of measles virus circulation in communities before clinical cases are recognized and can support public health responses to these re-emerging infectious diseases.

## Introduction

Measles, also known as rubeola, is a highly contagious viral illness caused by the measles virus (MV) (1,2). MV is a single-stranded RNA virus in the genus *Morbillivirus* and the family *Paramyxoviridae* (3). The virus primarily spreads through respiratory droplets and airborne transmission when an infected individual coughs or sneezes (4,5). The characteristic symptoms include a rash covering the body, a runny nose, fever, red watery eyes, small white spots inside the cheek areas, and coughing (6). Further exacerbation of the infection can lead to blindness, encephalitis, ear infections, pneumonia and other respiratory issues, diarrhea, and mortality (6). The introduction of the measles vaccine in 1963 dramatically reduced disease burden, however, measles remains endemic in some countries with continued outbreaks reported worldwide. In 2023, there were around 107,500 estimated deaths attributed to measles globally, mostly amongst unvaccinated or partially vaccinated children under the age of five (7,8).

On January 23, 2025, the Texas Department of State Health Services issued a health alert announcing two confirmed measles cases in Gaines County, Texas (9). As of April 4, 2025, this total has increased to 481 cases in 19 counties, mostly concentrated in South Plains and Panhandle region of Texas (10). There have been 56 total hospitalizations and two fatalities of unvaccinated children who lived in the original outbreak county (11). Approximately 33% of cases are in children under age 5, 40% in children aged 5 - 17, and 21% in adults. The ages of the remaining cases have not yet been confirmed (10). Of these cases, approximately 98% individuals had no documented doses of measles vaccinations prior to two weeks before symptom onset (10). In 2024, Texas was in the second lowest sextile of states with measles cases (1-9 cases), but by March 2025, it had the highest number of cases nationwide, exceeding the case count of the entire country from 2020 to 2024 (12).

Wastewater-based surveillance is an innovative approach for monitoring community-level outbreaks of infectious diseases, such as COVID-19 (13–17), influenza (18–21), and mpox (22– 25). Infected individuals can excrete viral particles into wastewater, regardless of whether they display clinical symptoms, thus allowing for a more accurate assessment of disease prevalence (26–29). Moreover, as viral shedding often begins prior to symptom onset (30–33), wastewater surveillance data can detect pathogen circulation before clinical cases are diagnosed, serving as an early warning system (34–38). This enables public health teams to better prepare medical infrastructure for any necessary care and implement timely prevention efforts (39–41). In this study, we investigated wastewater samples from two cities in the South Plains and Panhandle region of Texas to determine the presence of measles virus RNA in wastewater and track its temporal circulation in the community.

## Materials and Methods

### Wastewater sample collection

Raw influent wastewater samples were collected weekly from City A and City B in Texas from January 2, 2025, to March 17, 2025 (**Table S1**). These were 24-hour, composite samples collected from two wastewater treatment plants (WWTPs) in City A and City B, serving populations of 266,750 and 102,664 respectively (19). As of April 4, City A has reported over 30 active cases, while City B has reported none. Samples were collected and shipped overnight on ice in a secondary container to the laboratory in Houston, TX for analysis. Samples were processed on the day of receipt. There were 12 samples from City A and 10 samples from City B in total that were analyzed. Each sample was centrifuged into pellet and supernatant portions (3900 rpm for 10 mins), which was filtered through a 0.22 μm vacuum driven filtration system (Millipore, Cat. # SCGP00525) to remove any remaining cell detritus. The supernatants were then utilized to concentrate viral particles.

### Viral enrichment and extraction, and real-time PCR

The viral enrichment procedure was performed following previously tested methods (25,42,43). First, 15 mL of the filtrate was centrifuged (3900 rpm for 20 min) with the 30 kDa Amicon Ultra Centrifugal Filter (Sigma, Cat.# UFC9010) to 150 – 200 μL, which was used for RNA extraction via the QIAamp RNA Mini Kit (Qiagen, Cat.# 52906) following the manufacturer’s protocol. RNA was eluted with 100 μL of nuclease-free H_2_O. The eluted RNA was tested for measles virus using the Taqman™ Fast Virus 1-Step Master Mix (Bio-Rad, Cat.# 12010177) with the protocol: reverse transcription at 50°C for 10 minutes, initial denaturation at 95°C for 3 minutes, then 48 cycles of denature at 95°C for 10 seconds followed by an annealing/extending step at 60°C for 30 seconds. All primers and probes for measles were ordered from Integrated DNA Technologies.

We tested the samples with three molecular assays, MV-N3, MV-M1, and MV-M2, targeting the Nucleoprotein (N) and Matrix (M) genes, with M1 and M2 sharing identical primers but different probe sequences (**Table 1**). All probes were double-quenched; N3 and M2 assays utilized FAM-labeled probes, while the M1 assay used a HEX-labeled probe. For each assay, 8 technical replicates were conducted for each sample. Each PCR plate was run with 8 negative controls (nuclease free H2O). Cycle threshold (Ct) values were determined for each reaction, representing the number of amplification cycles required for the fluorescent signal to exceed the background threshold. Ct < 40 is considered positive detection (with secondary check on fluorescence growth curve). All wastewater samples were also tested for pepper mild mottle virus (PMMoV) as a reference control.

**Table 1.**
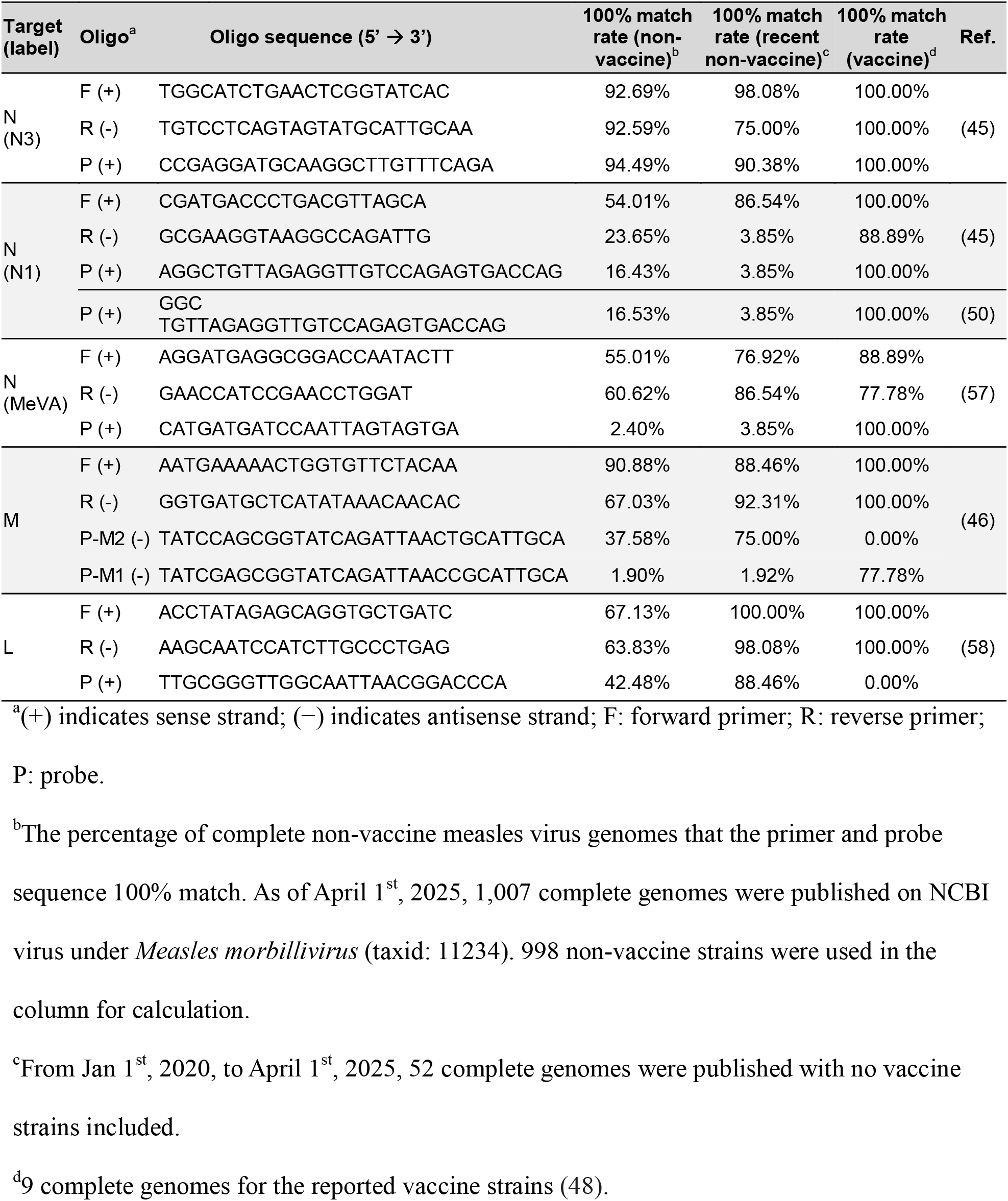
Perfect match rate of published primers and probes for measles detection.

### Sanger sequencing, alignment, and phylogenetic analysis

Positive RT-PCR products using M1 assay (168 bp) were further amplified with standard PCR using the same forward and reverse primers with Q5 high-fidelity DNA polymerase (New England Biolabs, Cat.#: M0544L). The amplicons were Sanger sequenced with both forward and reverse primers. The sequencing results were queried with NCBI BLASTn tool for species identification (Standard databases (nr etc.); and Highly similar sequences (megablast), Expected threshold: 0.05, Match/Mismatch Scores: 1, -2). For genotype comparison, complete genomic sequences of all available vaccine strains (n=9), along with representative sequences (from five different submitters) for major measles genotypes, including D8, B3, H1, and D4, were downloaded from NCBI GeneBank. Multiple sequence alignment was performed using MUSCLE in SnapGene (v7.1) to compare the sample sequences with reference genomes and assess phylogenetic relatedness. Multiple sequence alignment were performed using MUSCLE in SnapGene (version 7.1) to compare the wastewater sample sequences with those genomes and assess phylogenetic relatedness.

For a more comprehensive phylogenetic comparison, we downloaded all available complete genome sequences and associated metadata for the measles virus (*Measles morbillivirus*, taxid: 11234) from the NCBI Virus database as of April 1, 2025 (n = 1,007). Sequences without genotype information or collection date were excluded. The nine vaccine strains were manually labeled as ‘vaccine’ for easy visualization. The genomic sequences were truncated to match the length and position of our sanger sequencing results. All the downloaded sequences and our sanger sequencing results were then subjected to Nextstrain pipeline for phylogenetic analysis (44). The resultant phylogenetic tree was visualized with Auspice.

## Results

### Detection of measles viral RNA via three assays in the wastewater

We analyzed weekly wastewater samples collected from two cities in north Texas (City A and City B) from January 2, 2025, to March 17, 2025 (**Table S1**). To enhance detection sensitivity, we employed three previously validated assays targeting the measles virus genome: MV-N3 (45), MV-M1, and MV-M2 (46). Given the potentially low concentrations of measles virus RNA in wastewater due to limited community prevalence (47), we ran 8 replicates for each sample to maximize detection probability. PMMoV was tested in each sample with three replicates and showed consistent detections in each of the sample (**Figure S1**).

As shown in **Figure 1**, the RT-PCR results revealed varying detection patterns across sampling locations, dates, and assays. The heatmap displays the number of positive replicates (ranging from 1 to 4 out of 8) with corresponding Ct values. In City A, the first positive detection occurred on January 13 with the M2 assay (Ct 39.0), which is 10 days before the official outbreak announcement by the Texas Department State Health Services on January 23. The detection pattern in City A showed increasing consistency over time, with the strongest signals observed on February 10 and March 10, when all three assays detected viral RNA with multiple positive replicates. The N3 assay showed the most consistent detection in City A, yielding positive results in most samples from late January through March. In City B, measles virus RNA was detected as early as January 6 with the M1 assay showing 4 positive replicates (Ct 36.3), preceding the outbreak announcement by more than two weeks. The M1 and M2 assays showed concordant results for samples collected on January 13 and February 17, whereas the N3 assay yielded no positive results in City B throughout the study period. Across all positive samples, Ct values ranged from 35.4 to 39.8, indicating relatively low but detectable viral concentrations.

**Figure 1.**
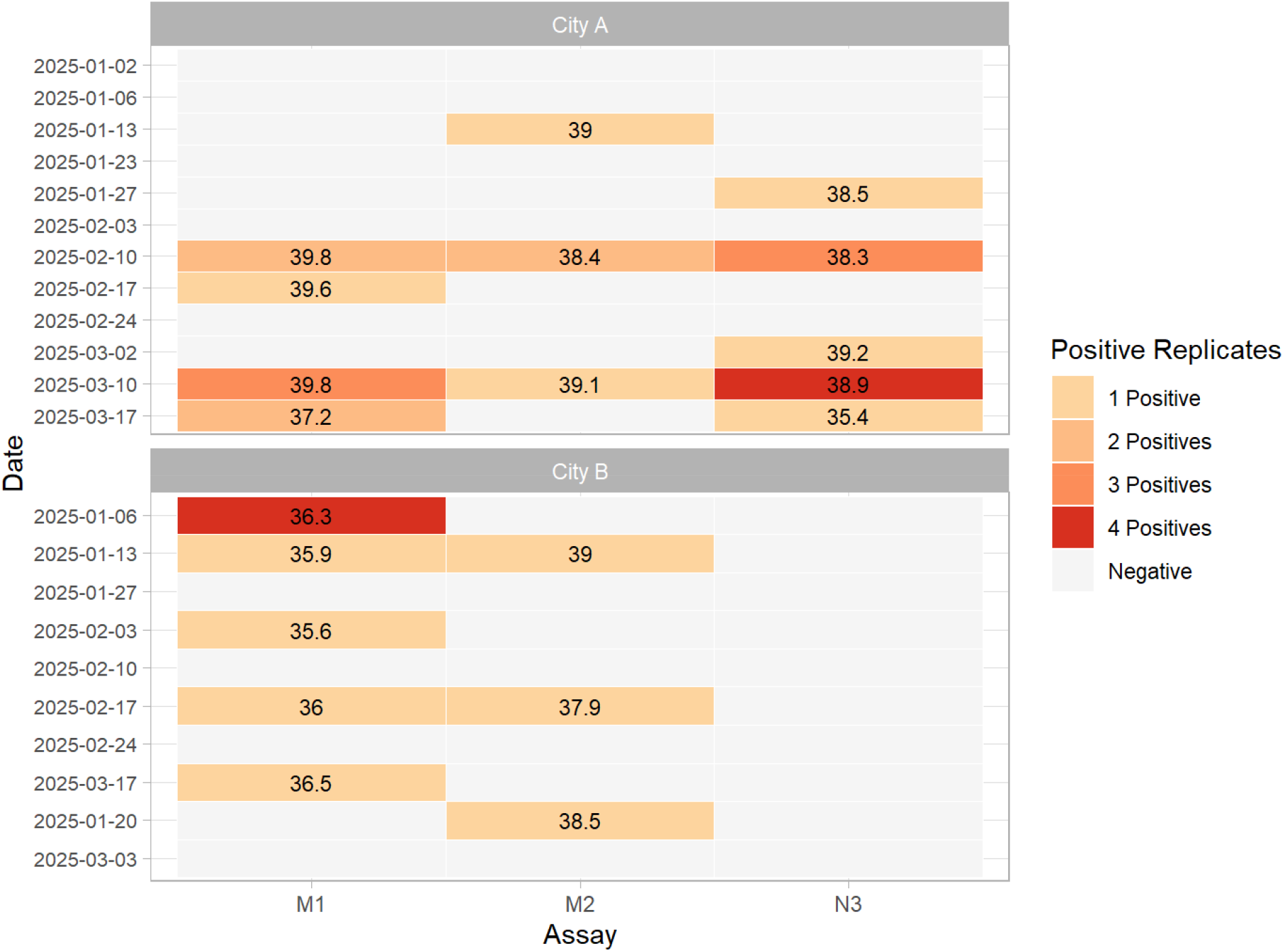
Detection of measles virus RNA in wastewater samples from two Texas cities (A and B). The heatmap shows the RT-PCR detection results with three different assays (M1, M2, and N3) for measles virus in wastewater samples from January 2 to March 17, 2025. Colors indicating the number of positive replicates (from 1 to 4) out of 8 total replicates per sample, and numbers representing Ct value or mean values.

These findings demonstrate that measles virus RNA was present in wastewater from both cities before the official outbreak announcement on January 23, confirming the utility of wastewater surveillance as an early warning system for detecting community transmission of measles virus.

### Confirmation of Measles Virus Sequence in wastewater samples

To validate the RT-PCR positive results and confirm measles virus detection, we sequenced the amplicons using both forward and reverse primers. Gel electrophoresis confirmed the expected band size (close to 200 bp). BLASTn analysis of the sequencing data showed that all these sequences aligned to Measles virus genotype D8, with an average of 96.78% (84.62%-100%) sequence identity.

To distinguish between vaccine and wild-type strains and validate the genotype classification, we performed a multiple sequence alignment using representative genomes from major measles virus genotypes, including the currently circulating genotypes D8 and B3, historically prevalent genotypes H1 and D4, and all nine documented vaccine strains (48). For each genotype, sequences from different submitters were selected to minimize sampling bias and reduce the risk of submitter-specific artifact. As shown in **Figure 2**, the wastewater-derived sequences were identical to the D8 references within the target region and displayed distinct mutation signatures that differentiate them from vaccine strains and other genotypes (varied nucleotides were highlighted). These shared substitution patterns support the classification of the measles RNA detected in wastewater samples as genotype D8.

**Figure 2.**
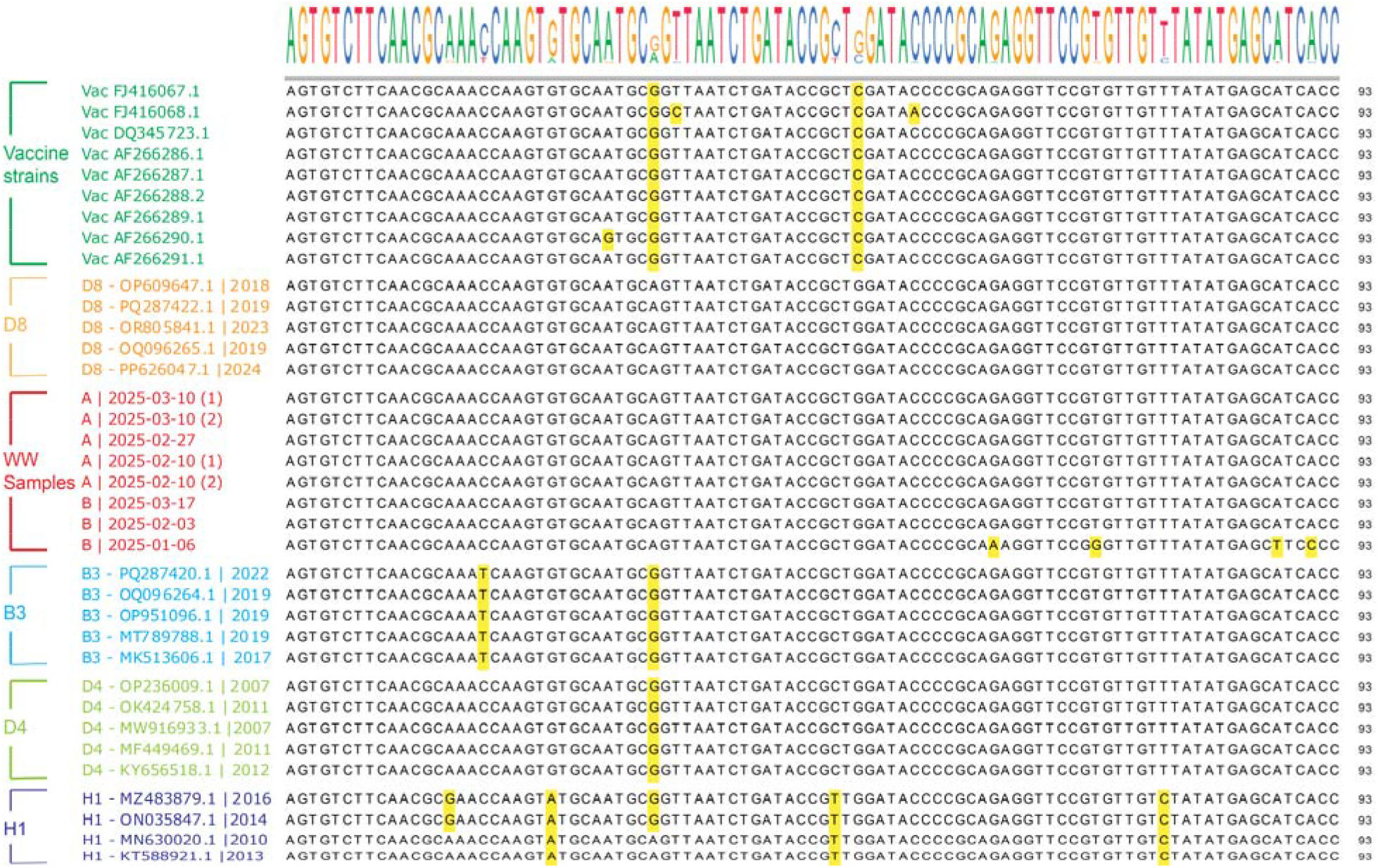
Multiple sequence alignment of measles virus sequences detected in wastewater with reference genotypes. Alignment showing nucleotide variations in the amplified region from wastewater-derived sequences compared with representative sequences from major measles virus genotypes (including D8, B3, H1, D4) and vaccine strains. Genotype, accession number, and sampling year for each sequence were indicated. Variant nucleotides are highlighted in yellow. Wastewater-derived sequences show identical patterns to D8 references, with distinct mutation profiles that differentiate them from vaccine strains and other genotypes.

For a broader phylogenetic context, we constructed a maximum-likelihood tree using 1,007 complete measles virus genomes obtained from NCBI Virus (downloaded as of April 1, 2025), after excluding records lacking genotype information or collection date. In line with current surveillance data, genotypes D8 and B3 are still in global circulation (**Figure 3A**). The sequenced wastewater samples clustered within the D8 clade, consistent with both the alignment results and current epidemiological trends. Similar clustering pattern was also found when only using the USA dataset (**Figure 3B**). Together, the sequence alignment and phylogenetic analyses confirm that the detected measles virus RNA in wastewater samples are true positives and belong to genotype D8.

**Figure 3.**
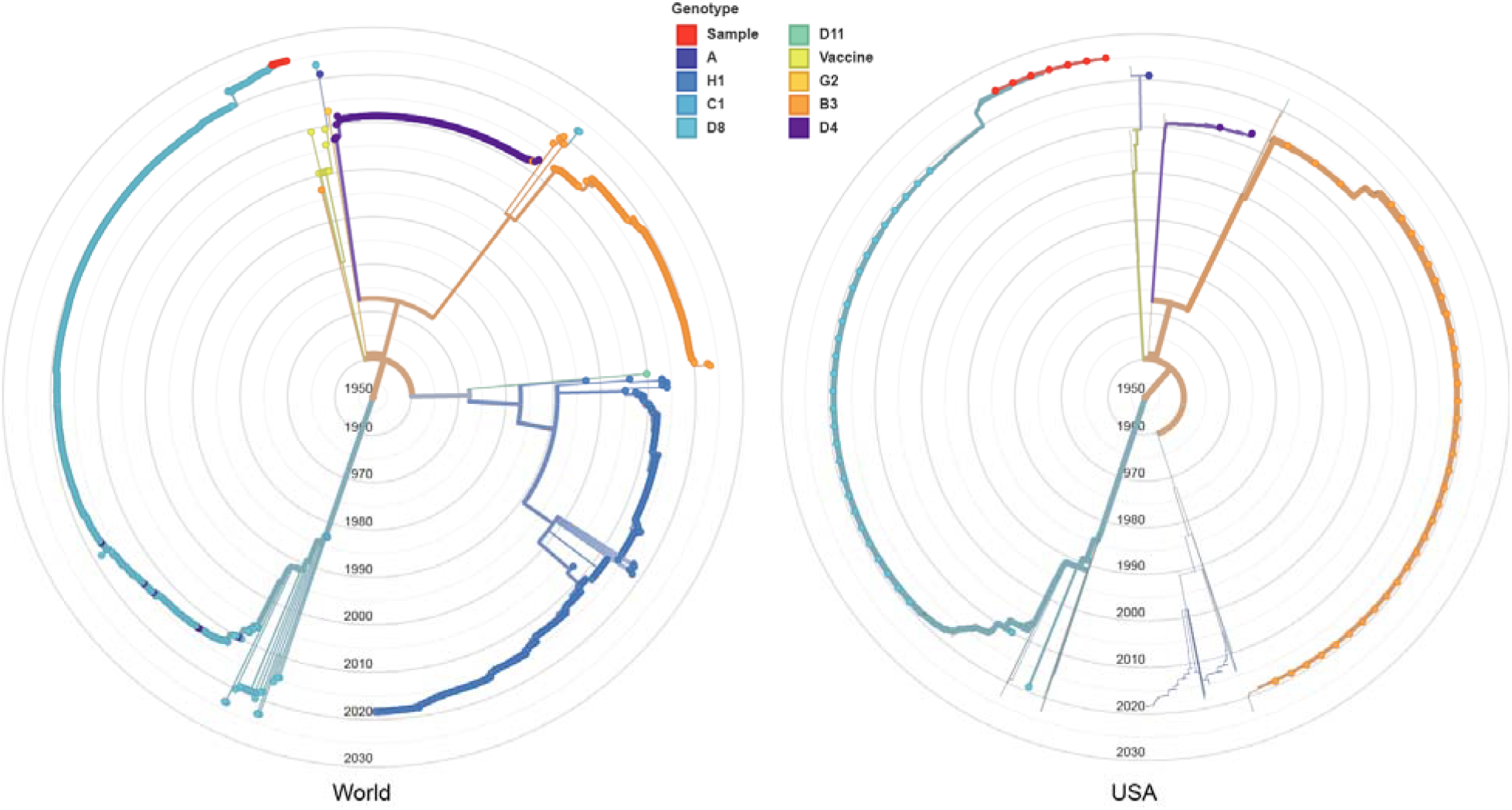
Phylogenetic analysis of measles virus sequences detected in wastewater. **(A)** Maximum-likelihood tree constructed using 1,007 complete measles virus genomes from NCBI Virus database (as of April 1, 2025), showing global distribution of measles genotypes. **(B)** Phylogenetic analysis focused on USA measles virus sequences. Both trees display temporal evolution from the center (older isolates, 1960s) to the periphery (recent isolates, 2020s). Different genotypes are color-coded as indicated in the legend, with vaccine strains in yellow, and genotypes A, B3, C1, D4, D8, D11, G2, and H1 in their respective colors. The wastewater-derived sequences from this study (red) align with recently circulating D8 genotype (light blue). Time-resolved phylogenetic trees were built with Nextstrain and visualized in radial format with Auspice.

## Discussion

Our study demonstrates the effectiveness of wastewater-based surveillance as a timely warning system for measles outbreaks, with viral RNA detected and sequence confirmed in City B as early as January 6, predating the official outbreak announcement by more than two weeks. Through integration of RT-PCR detection with multiple assays, amplicon sequencing, and phylogenetic analysis, we provide evidence of measles virus genotype D8 circulation in community wastewater during this active outbreak in Texas. These findings establish wastewater surveillance as a valuable complementary approach to traditional clinical surveillance and to monitoring ongoing outbreaks.

Wastewater surveillance for measles is an emerging approach with growing evidence supporting its feasibility. Recent modelling work by Bibby et al suggest that achieving 50% probability of detection requires approximately 78 cases per 100,000 people with a process limit of detection of 1000 genome copies per liter (47). This generally aligns with a recent Switzerland study that found positive signals in wastewater when 21 confirmed cases occurred in a week within a catchment area of 240,000 people, with no detection when cases fell below this threshold (49). Two most recent studies have successfully detected measles virus genotype A in Canadian wastewater and genotype D8 in Belgium wastewater (50,51). Our findings further extend this work by demonstrating measles virus detection preceding clinical outbreak recognition and confirming the outbreak strain through sequencing and phylogenetic analysis. While City A reported over 30 confirmed measles cases (during this study period) starting in late January, City B had no officially reported cases despite detection of measles virus RNA in its wastewater as early as January 6. The positive signals in City B’s wastewater likely represent unrecognized viral circulation that could be attributed to subclinical infections, asymptomatic cases, or transient travelers (52,53). This finding may also suggest the need for enhanced clinical monitoring in the area.

While our findings confirm the value of wastewater surveillance for measles detection, several areas need further development. First, determining precisely how far in advance wastewater signals can detect viral circulation before clinical cases emerge is crucial for maximizing the early warning benefits of this approach. In our study, detection preceded official case announcements by approximately two weeks, but systematically quantifying this lead time across different outbreaks and settings would be valuable for public health response planning. Second, establishing quantitative relationships between wastewater viral concentrations and community case numbers would enhance interpretability of surveillance data. Our study reported the Ct values, but did not attempt to correlate these with case counts due to limitations in case data granularity at the sewershed level. Third, developing mechanistic mathematical models to translate viral concentrations into prevalence estimates would strengthen the epidemiological utility of wastewater data, as demonstrated in earlier studies on other viral pathogens (26,54). Last but not the least, the selection of appropriate molecular assays is a critical factor influencing detection success in our study based on our experience with other viruses (25,55). The differential performance of assays between cities prompted us to conduct a comprehensive mismatch analysis of five commonly used measles RT-PCR assays (**Table 1**). This analysis revealed that the N3 assay serves effectively as a pan-measles detection tool with high match rates across all strain types (>90%), while the M assay offers strain differentiation capabilities through its dual probe design. Interestingly, while the M1 probe showed high theoretical specificity for vaccine strains (77.78% match rate) and M2 for wild-type strains (75% match rate for recent strains), both detected samples in our study that were later confirmed as wild-type D8 through sequencing. This highlights the need for performance verification in real-world environmental samples. Based on the results in **Table 1**, N3 has the best matching for broad measles surveillance, and M2 or the L assay show optimal performance for wild-type strain detection.

In conclusion, this study highlights wastewater-based epidemiology as a valuable approach for detecting measles outbreaks. As measles continues to pose local and global public health challenges due to vaccination hesitancy and program disruptions (1,56), incorporating wastewater surveillance into routine monitoring programs could provide earlier warning signals and support more timely public health responses to this re-emerging infectious disease.

## Data Availability

All data are provided in this work.

## Dada availability

All data are provided in this work.

## Acknowledgements

We thank the staff of the WWTPs in the respective cities for collecting the wastewater samples used in the study. This work is supported by the National Science Foundation (DMS-2421257) and the Texas Epidemic Public Health Institute (TEPHI).

## Supplemental Figures and Table

**Table S1:** Wastewater Sample Information.

| City | Sampling Date | Flow Rate (million gallons per day) | pH | Collection Temp | Sampling Day of the Week |
| --- | --- | --- | --- | --- | --- |
| City A | 1/2/2025 | 19.64 | 7.54 | 18.72 | Thursday |
| City A | 1/6/2025 | 19.33 | 7.57 | 17.8 | Monday |
| City A | 1/13/2025 | 19.33 | 7.57 | 22.4 | Monday |
| City A | 1/23/2025 | 18.79 | 7.48 | 18.1 | Thursday |
| City A | 1/27/2025 | 19.3 | 6.5 - 7.0 | 18.1 | Monday |
| City A | 2/3/2025 | 18.6 | 6.5 - 7.0 | 19.3 | Monday |
| City A | 2/10/2025 | 19.02 | 6.5 - 7.0 | 8.9 | Monday |
| City A | 2/17/2025 | 18.76 | 6.5 - 7.0 | 8.1 | Monday |
| City A | 2/24/2025 | 18.96 | 6.5 - 7.0 | 8.4 | Monday |
| City A | 3/2/2025 | 19.16 | 6.5 - 7.0 | 8.6 | Monday |
| City A | 3/10/2025 | 19.25 | 6.5 - 7.0 | 19.3 | Monday |
| City A | 3/17/2025 | 17.96 | 6.5 - 7.0 | 19.1 | Monday |
| City B | 1/6/2025 | 7.07 | 9.14 | 25 | Monday |
| City B | 1/13/2025 | 6.2 | 7.36 | 15.85 | Monday |
| City B | 1/20/2025 | 8.17 | 6.91 | 25 | Wednesday |
| City B | 1/27/2025 | 5.97 | 7.34 | 15.15 | Monday |
| City B | 2/3/2025 | 8.86 | 7.04 | 17.78 | Monday |
| City B | 2/10/2025 | 7.41 | 7.04 | 13.51 | Monday |
| City B | 2/17/2025 | 8.43 | 7.02 | 13.8 | Monday |
| City B | 2/24/2025 | 8.27 | 7.13 | 13.79 | Monday |
| City B | 3/3/2025 | 11.2 | 7.3 | 15.47 | Monday |
| City B | 3/17/2025 | 8.49 | 6.97 | 14.04 | Monday |

**Figure S1:**
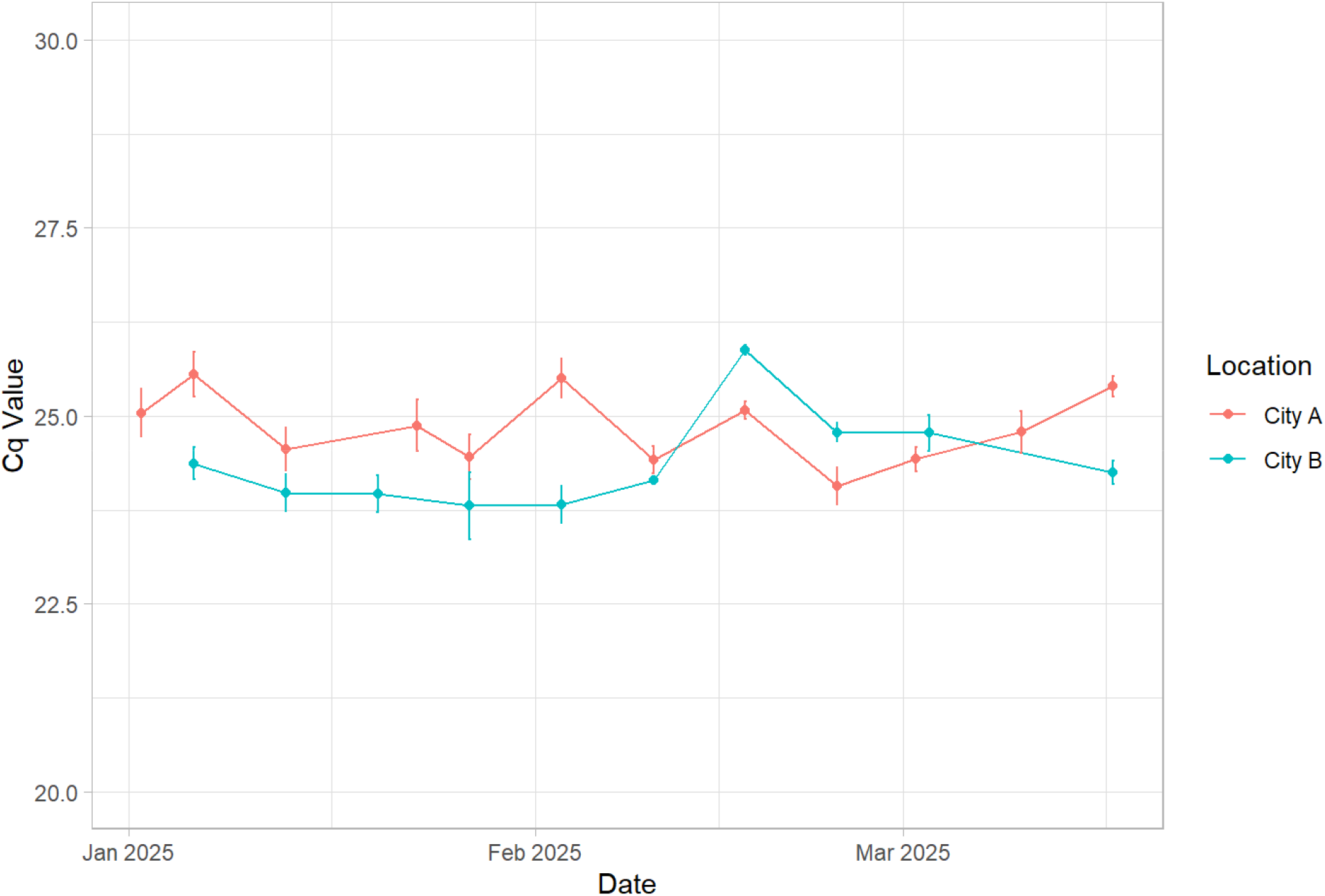
PMMoV Ct values for all samples tested in this study as an internal reference for the wastewater.

